# Replicating Global Brain Connectivity as an Imaging Marker for Depression – Influence of Preprocessing Strategies and Randomized Placebo-Controlled Ketamine Treatment

**DOI:** 10.1101/19010504

**Authors:** Christoph Kraus, Anahit Mkrtchian, Bashkim Kadriu, Allison C. Nugent, Carlos A. Zarate, Jennifer W. Evans

## Abstract

Major depressive disorder (MDD) is associated with altered global brain connectivity (GBC), as assessed via resting state functional magnetic resonance imaging (rsfMRI). Previous studies found that antidepressant treatment with ketamine normalized aberrant GBC changes in the prefrontal and cingulate cortices, warranting further investigations of GBC as a putative imaging marker. However, the results were only obtained via global signal regression (GSR). This study is an independent replication of that analysis using a separate dataset. GBC was analyzed in 28 individuals with MDD and 22 healthy controls (HCs) at baseline, post-placebo, and post-ketamine. To investigate the effects of preprocessing, three distinct pipelines were used: 1) regression of white matter (WM)/cerebrospinal fluid (CSF) signals only (BASE); 2) WM/CSF+GSR (GSR); and 3) WM/CSF+physiological parameter regression (PHYSIO). Compared to PHYSIO and BASE regression, GSR reduced Fisher Z-scores (Fz-scores) in large clusters. PHYSIO did not resemble GBC preprocessed with GSR (GBCr). Reduced GBCr was observed in individuals with MDD at baseline in the anterior and medial cingulate cortices, as well as in the prefrontal cortex. Significant results were only found with GSR. Ketamine had no effect compared to baseline or placebo in either group. These results concur with several studies that used GSR to study GBC. Altered GBCr was observed in the cingulate and prefrontal cortices, but ketamine treatment had no effect. Further investigations are warranted into disease-specific components of global fMRI signals that may drive these results and of GBCr as a potential imaging marker in MDD.

## Introduction

In contrast to other medical diseases, no objective biomarkers exist for major depressive disorder (MDD). In this context, functional MRI (fMRI)—which has greatly advanced our understanding of the aberrant neural circuits associated with MDD—may help researchers objectively quantify psychiatric disorders [1]. In particular, resting-state connectivity analyses that capture several minutes of brain activity at ‘rest’ are attractive as imaging markers, given that MRI is widely available and resting state scans are easier to record than task-based fMRI. However, resting state connectivity demands elaborate preprocessing to eliminate spurious signals such as motion-related artifacts and vascular or other physiological non-neuronal signals [1-3]. fMRI studies of individuals with MDD have reported dysconnectivity in networks involved with attention, emotion processing, salience, and goal-directed behavior [4-6]. These results overlap with MRI findings of reduced gray matter (GM) volumes and white matter (WM) connectivity in individuals with MDD [7-9]. While resting state fMRI (rsfMRI) connectivity measures are thus good candidate imaging markers for MDD, essential preprocessing and analysis procedures have nevertheless not been well established and need validation.

Global brain connectivity (GBC), also known as functional connectivity strength, is a correlation-based connectivity approach that has been proposed as an imaging marker for several psychiatric disorders [10-12]. GBC yields a three-dimensional correlation map for each fMRI scan [10]. It is calculated by correlating the time series of every GM voxel with every other GM voxel, transforming correlations to Fisher Z-scores (Fz-scores), and averaging these. While GBC assumes unweighted, linear correlations and does not account for spatial autocorrelations inherent in rsfMRI data, which could inflate Z-scores [13], correlation coefficients nevertheless yield nondirectional information on highly connected network hubs [10,14]. In several recent studies, GBC revealed dysconnectivity in MDD with very large effect sizes in the frontal cortex, posterior cingulate cortex, and cerebellum [11,12,15]. After treatment with the rapid-acting antidepressant ketamine, aberrant connectivity returned towards values observed in healthy participants [11,12].

Although these prior results were notable, group differences between individuals with MDD and healthy controls (HCs) were only observed after preprocessing fMRI data with global signal regression (GSR), as were changes following ketamine treatment [11,12,15,16]. GSR is a somewhat controversial procedure for cleaning fMRI signals, as it mathematically alters functional interactions by introducing negative correlations [17], downweights voxels with large net activity [18], and could induce artificial group differences [19,20]. However, the global signal (GS) contains spurious noise such as residual motion artifacts and physiological signals such as vascular and respiratory signals that are better removed [2].

To validate GBC preprocessed using GSR (GBCr) as a potential imaging marker for MDD and antidepressant treatment, the effects of GSR as a preprocessing step in GBC analyses should be better understood. This study sought to independently replicate the finding of disrupted GBCr in previous studies of individuals with MDD by recreating the preprocessing steps reported by the earlier studies [11,12]. The study also sought to examine the specific effects of GSR on GBC by comparing three common preprocessing strategies: 1) regression of WM/cerebrospinal fluid (CSF) signals only (hereafter referred to as BASE); 2) regression of WM/CSF signals plus regression of the total GM GS (GSR, which was used in previous studies [11,12]); and 3) WM/CSF plus physiological parameter regression (hereafter referred to as PHYSIO). Given that the GS contains physiological components, the effects of PHYSIO regression on GBC was hypothesized to be similar to GSR.

## Materials and Methods

### Participants and study design

Participants included in this study were part of a larger protocol at the National Institute of Mental Health (NIMH) in Bethesda, Maryland, USA (NCT00088699, National Institutes of Health (NIH) Protocol No. 04-M-0222). All participants provided written informed consent after oral explanation of study procedures, which were approved by the NIH Combined Central Nervous System Institutional Review Board. Independent analyses and results from these studies have previously been published [6,21-24].

All included participants were between 18 and 65 years old and underwent a double-blind, placebo-controlled, crossover study with intravenous infusion of ketamine hydrochloride (0.5 mg/kg) or saline two weeks apart in randomized order, to minimize potential carryover effects. Additional details, including demographic characteristics and inclusion/exclusion criteria for each particular substudy, are available in the Supplementary Methods. Thirty-three individuals with MDD and 25 HCs underwent MRI scans. Of these, 30 participants with MDD and 24 HCs had complete datasets including physiological data, and 28 MDD participants and 22 HCs were available for statistical analysis after censoring (see Table 1 and Supplementary Table S1). All MDD and HC participants underwent a baseline scan and a scan on Day 2 or 3 after ketamine or placebo infusion. Additional imaging details are available in the Supplementary Methods.

**Table 1.**
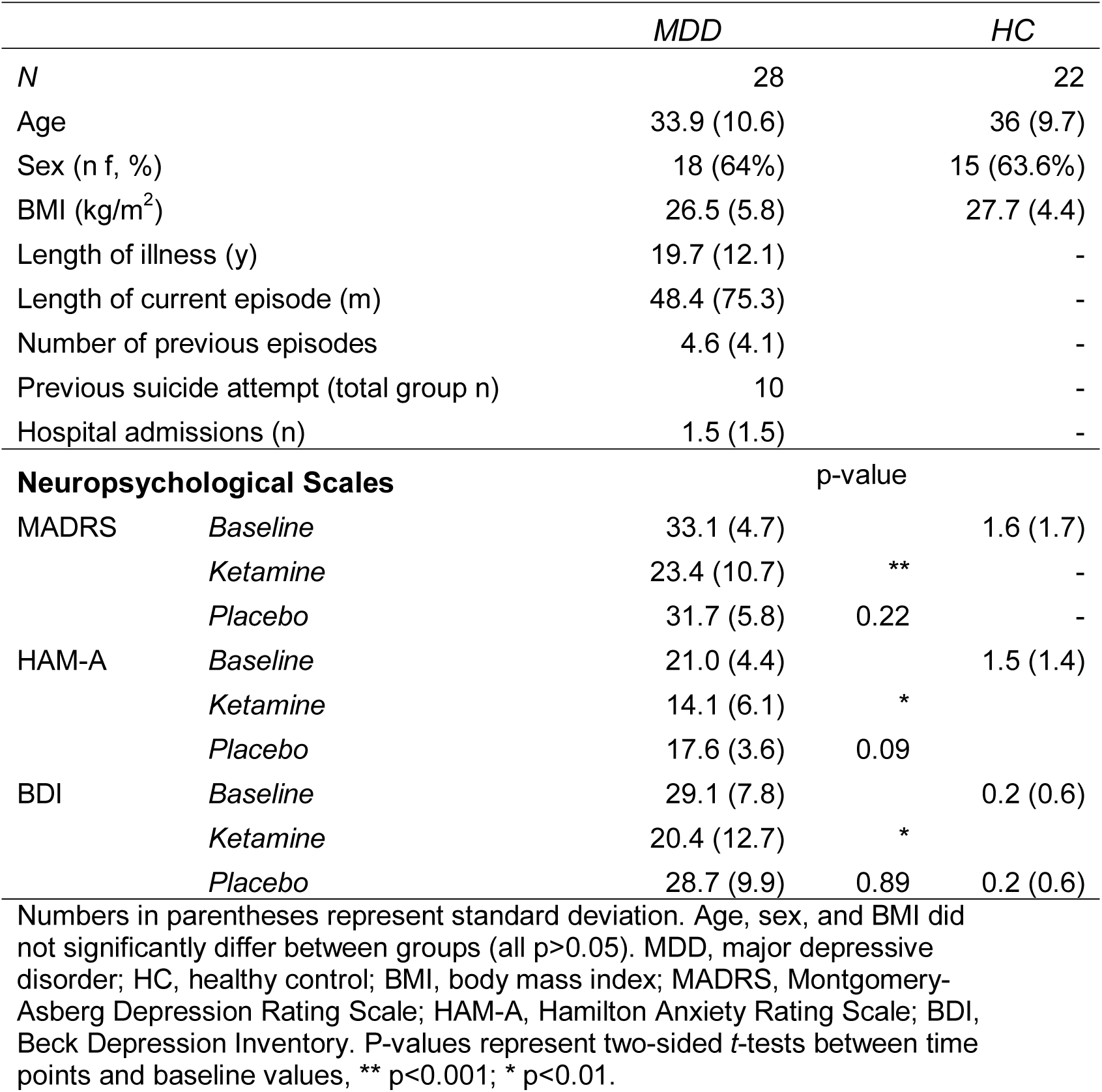
Participant Characteristics (maximal *N*, analyzed sample)

MDD participants were diagnosed using the Structured Clinical Interview for DSM-IV (SCID-I, patient version), and all were currently experiencing a major depressive episode. All MDD participants were admitted to our inpatient unit and were treatment-resistant, defined as having failed to respond to at least one previous adequate antidepressant trial, assessed via the Antidepressant Treatment History Form [25]. Severity of the current episode was measured using the Montgomery-Åsberg Depression Rating Scale (MADRS); all participants had a total score at screening and prior to every infusion of ≥ 20. Psychometric mood assessments were conducted with the MADRS, Beck Depression Inventory (BDI), and Hamilton Anxiety Rating Scale (HAM-A) before, during, and after each infusion, as previously described [6].

### Preprocessing procedures and global brain connectivity calculation

All preprocessing steps are illustrated in Supplementary Figure S1, and detailed descriptions of rsfMRI preprocessing procedures can be found in the Supplementary Methods.

Structural MRIs for each baseline scan were segmented into GM, WM, and CSF with FreeSurfer version 5.3 (see Supplementary Figure S2). Preprocessing of all rsfMRI data was conducted with AFNI version 19.0.09 [26]. To investigate the effects of preprocessing, three distinct pipelines were used: 1) regression of WM/CSF signals only (BASE); 2) regression of WM/CSF signals plus regression of the total GM GS (GSR); and 3) WM/CSF plus physiological parameter regression (PHYSIO) in AFNI’s ANATICOR [27].

As with previous studies investigating GBC in individuals with MDD [11,12], all rsfMRI preprocessing for the three preprocessing pipelines was conducted in native space with AFNI’s 3dTcorrMap program within each individual participant’s total GM, including cerebellum and basal ganglia. Each participant’s GBC map was then transformed to standardized space (Montreal Neurological Institute, MNI_caez_N27) only for second-level statistics. During normalization, GBC maps were resampled to a final voxel size of 3.5 mm isotropic (voxel volume: 42.875 mm^3^). As with previous studies of this issue [11,12], and in order to compare results, intra-prefrontal cortex (PFC) GBC was also calculated. This was done using identical procedures for all three of the preprocessing pipelines used for whole brain GBC analysis, but GBC calculations were restricted to the PFC with a PFC GM-mask (see Supplementary Table S2).

### Statistics

#### Comparing preprocessing pipelines

The variance explained (R^2^) by regressors in each pipeline was investigated in order to compare their impact across groups and time points. R^2^ was calculated for each regressor in total GM for each scan as well as in voxel-wise R^2^ maps, and differences were compared using linear mixed models. Detailed methods describing comparisons of the R^2^ for each pipeline-specific regressor can be found in the Supplementary Methods.

Baseline GBC maps created using the three pipelines were then compared to assess how they differed. We hypothesized that GBC maps generated with PHYSIO regression would resemble those created using GSR. This was based on evidence demonstrating that cardiac and respiratory response functions can be derived from the GS [28]. For comparison, the same 22 HCs for whom GBC baseline maps were available in all three pipelines were also included. Analysis of variance (ANOVA) and post-hoc *t-*tests were conducted with 3dMVM, with pipeline as the factor of interest.

Results were corrected for family-wise error (FWE) at p<0.05 (two-tailed) by using the AFNI routines 3dFWHMx and 3dClustSim with an initial threshold of p=0.001 (cluster size>7).

The statistical methods used to analyze clinical effects can be found in the Supplementary Methods.

### Group comparisons between baseline, ketamine, and placebo scans

Differences in whole brain GBC between baseline, ketamine, and placebo scans within and between groups (MDD and HC) were calculated independently for each preprocessing pipeline using the AFNI routine 3dLME. Each model had main effects for group and scan as well as their interaction as fixed effects, and participant was included as a random effect. Post-hoc contrasts between groups (MDD vs. HC) were calculated at each time point (baseline, ketamine, placebo), as were contrasts between each pair of time points (ketamine-baseline, placebo-baseline, ketamine-placebo). The same contrasts were also calculated within each group, which tested effects separately in HC and MDD participants. To investigate the effects of age, sex, and body mass index (BMI), the mixed model controlling for these variables was also repeated. Group GBC maps were corrected by estimating a Gaussian-shaped autocorrelation function with FWHMx with FWE-corrected cluster thresholds of p<0.05; these were calculated with 3dFWHMx and 3dClustSim with an initial threshold of p=0.001 (cluster size>8). To allow our results to be compared with previous studies [11,12], an initial threshold of p<0.01 was used, which resulted in a cluster size of 23 for p<0.05, FWE-corrected. Identical statistics were repeated with intra-PFC GBC maps.

## Results

### Ketamine’s clinical effects

In individuals with MDD, a significant effect of time was observed on MADRS (F_2,69_=12.58, p<0.001), BDI (F_2,60_=6, p=0.003), and HAM-A (F_2,67_=7.42, p=0.001) scores. Compared to placebo, ketamine treatment significantly reduced MADRS (t_35_=- 3.32, p=0.002), BDI (t_43_=-2.63, p=0.012), and HAM-A (t_38_=-2.4, p=0.022) scores compared to baseline values.

### Comparing GBC measures obtained with different preprocessing strategies

We compared the R^2^ from all three preprocessing-specific regressors: BASE, GSR, and PHYSIO. In the baseline scans for the 22 HCs, a significant main effect of R^2^ was observed for every regressor (F_2,426_=278.6, p<0.001); BASE regression explained significantly more variance in all GM voxels than GSR (t_41589_=255.8, p<0.001, false discovery rate (FDR)-corrected) or PHYSIO regression (t_41591_=17.5, p<0.001, FDR-corrected, see Supplementary Figure S3C).

When R^2^ raw values for each regressor were compared between scans and groups with mixed models, no significant main effect was observed for group (Supplementary Figure S3 and Supplementary Table S3). However, a significant main effect was observed for scan (baseline, ketamine, placebo; χ^2^=6.21, df=2, p=0.045) for GSR (χ^2^=8.09, df=2, p=0.017), and this effect was not present for PHYSIO regression (χ^2^=3.58, df=2, p=0.167). Post-hoc tests found significantly lower R^2^ values using BASE regression in MDD participants between both ketamine (t_92_=2.5, p=0.014, uncorrected) and placebo (t_95_=2.58, p=0.012, uncorrected) scans compared to baseline. In individuals with MDD, GS accounted for significantly lower R^2^ values between baseline and placebo scans (t_95_=2.68, p=0.009, uncorrected; see Supplementary Table S4).

Analysis of spatial maps corresponding to the BASE, GSR, and PHYSIO pipelines yielded significant effects of time point for R^2^ maps corresponding to BASE regression in the right lingual cortex, right temporoparietal junction, and right inferior temporal cortex (F-tests: 9-9.6, all p<0.05, FWE-corrected, Supplementary Figure S4). For GSR, significant main effects were observed for group in the right lingual cortex spreading to the cerebellum (F=15, p<0.05, FWE-corrected). Significant main effects for scan were also noted in the left supramarginal cortex and the left precuneus (F=8.9-9.1, p<0.05, FWE-corrected). No significant main effects or interactions were seen for group or scan with PHYSIO regression (all p>0.05, FWE-corrected). Supplementary Figure S4 provides a spatial map of R^2^ values in 22 HCs indicating spatial overlap of GSR R^2^ with the venous system (for additional details, see Supplementary Results, Supplementary Figure S5, and Supplementary Table S5).

The ANOVA for the GBC of the baseline scan between all three preprocessing pipelines in 22 HCs demonstrated a significant main effect of pipeline (p<0.05, FWE-corrected). Brain-wide reductions in Fz-scores were observed after GSR compared to BASE regression (cluster spreading bilaterally from cerebellum to anterior with a volume of 316,090 mm^3^, p<0.05, FWE-corrected, see Figure 1A-C and Supplementary Table S3). GSR resulted in a numerically robust reduction (*d*=1.65) of Fz-scores and introduced negative numbers, especially in terminal fields along the anterior-posterior axis (Figure 1A, 1B). In contrast, no significant differences were observed between PHYSIO and BASE regression only (all uncorrected p>0.001). A similar cluster pattern was observed when comparing GSR with PHYSIO regression (data not shown).

**Figure 1.**
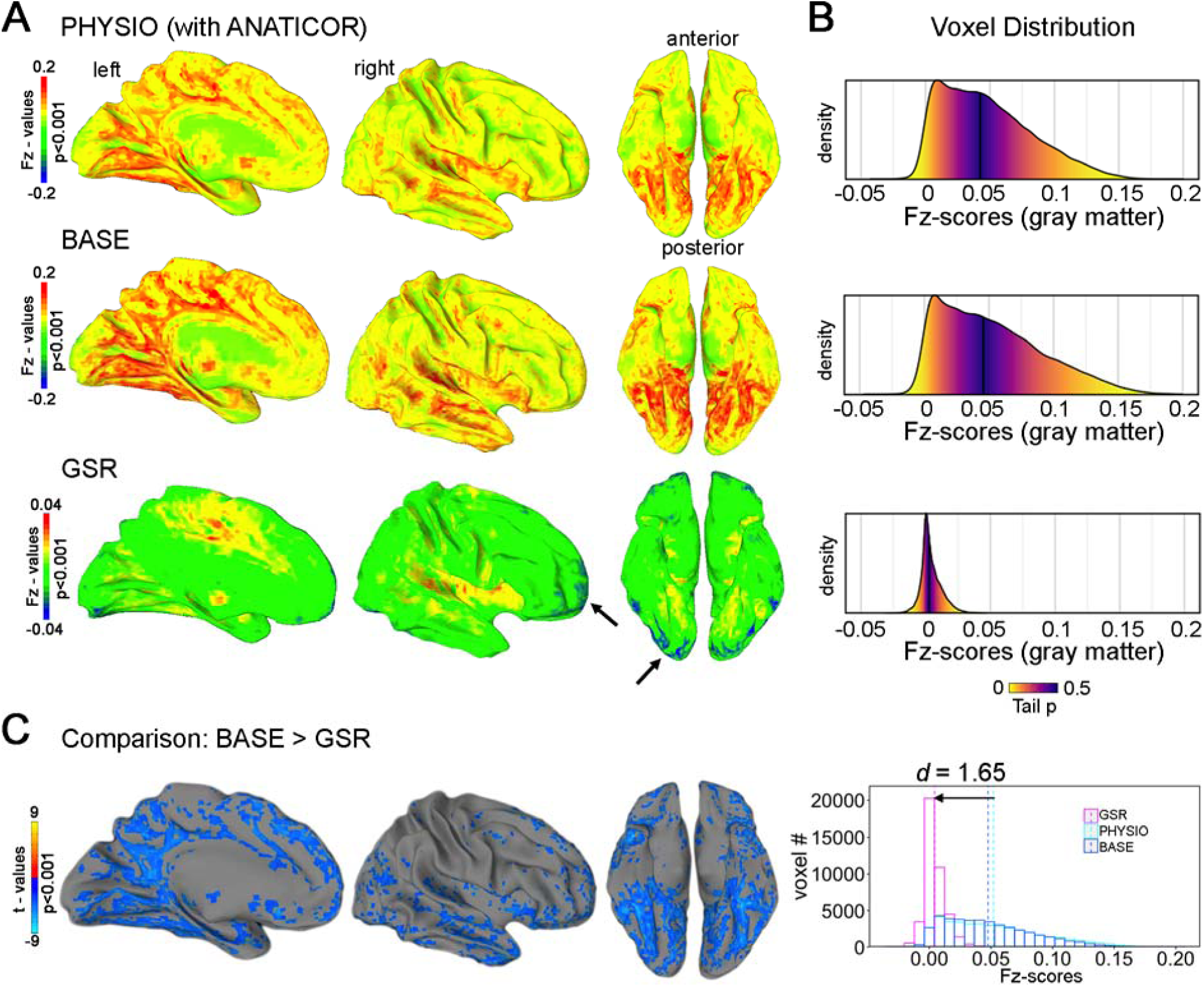
Comparison of the three preprocessing strategies on measures of global brain connectivity (GBC) in 22 healthy controls (HCs) at baseline. **A**, Unthresholded GBC maps (group average) with WM/CSF regression (BASE) resembled those with WM/CSF regression+physiological parameter regression (PHYSIO). GBC maps after WM/CSF+global signal regression (GSR) yielded peaks around the basal ganglia, insulae, and cingulate cortices. Negative Fisher transformed Z-scores (Fz-scores) were introduced in terminal fields along the anterior-posterior axis. **B**, Absolute Fz-score distributions corresponding to (**A**). Fz-scores centered around zero after GSR; color-coding represents tail probabilities around the median (dark line). **C**, Comparison between BASE regression and GSR GBC maps yielded strong brain-wide reductions in Fz-scores with an emphasis on posterior-basal regions. Histograms depict total gray matter (GM) Fz-scores in 22 HCs at baseline; lines represent means.

### Whole brain GBC results across all pipelines

Using the three common preprocessing strategies—BASE, GSR, and PHYSIO regression—significant GBC changes were observed only after GSR, both with an initial cluster-defining threshold of p<0.001 as well as with a more lenient p-value of p<0.01 (both corrected for FWE at p<0.05). A significant main effect was found for group across all time points in the left cerebellar lobules VII-VIII (F=18.2, p<0.001, FWE-corrected) as well as in the bilateral medial cingulate cortex (MCC, F=18.3, p<0.001, FWE-corrected; see Figure 2 and Supplementary Table S6). At baseline, individuals with MDD exhibited significantly lower GBCr compared to HCs in the bilateral MCC (Z=3.9, p<0.001, FWE-corrected, *d*=1.76; see Figure 2A, 2B). Controlling for age, sex, and BMI did not change these results (see Supplementary Table S6). As in previous studies [11,12], the p-value was lowered to <0.01; significant main effects were again observed in the cerebellum and MCC as well as in the right middle temporal cortex. At baseline, GBCr was significantly reduced bilaterally in the anterior cingulate cortex (ACC) spreading to the superior medial frontal cortices (Z=2.9, p<0.01, FWE-corrected, Figure 2B). No significant main effect was observed for scan (baseline, ketamine, placebo) or interaction between group and scan (all p>0.05, FWE-corrected). In addition, reduced baseline Fz-scores in participants with MDD in the MCC cluster were not detected with BASE or PHYSIO regression (Figure 2C, bottom).

**Figure 2.**
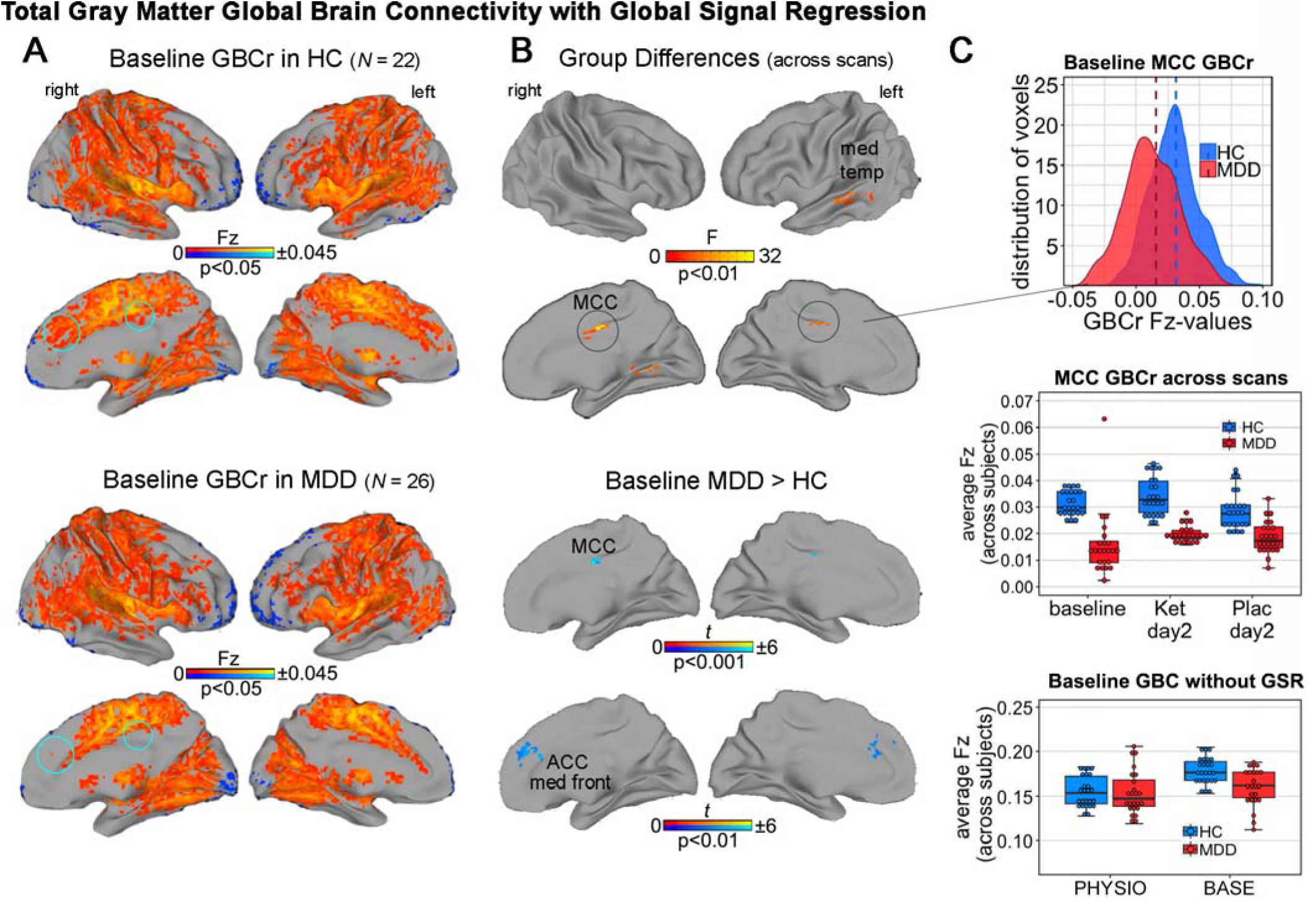
Total gray matter (GM) global connectivity with global signal regression (GSR). Reductions in total GM global brain connectivity (GBC) observed with white matter (WM)/ cerebrospinal fluid (CSF)+GSR (GBCr) in individuals with major depressive disorder (MDD). No significant results were observed with the other preprocessing pipelines (all p>0.05, FWE-corrected). **A**, Average baseline connectivity in both groups (p<0.05, uncorrected). Blue circles indicate areas demonstrating significant reductions in individuals with MDD compared to healthy controls (HCs). **B**, Top: main effect of group across all baseline, post-ketamine, and post-placebo scans. Bottom: reduced connectivity in individuals with MDD compared to HCs in the baseline scan in the medial cingulate cortex (MCC) and anterior cingulate cortex (ACC) as well as in the medial frontal cortex (*t*-tests, p<0.001 and p<0.01, FWE-corrected at p<0.05). **C**, Reduced GBCr values in individuals with MDD at baseline (top) were also observed across scans, leading to the significant main effect of group across scans depicted in (**B**). Reduced GBC values in the MCC were not observed with the other preprocessing strategies (bottom). No main effect of drug or post-hoc GBCr differences between ketamine, placebo, or each versus baseline were found within the HC or MDD groups (all p>0.05, FWE-corrected). For statistical results see Supplementary Table S6.

### Intra-PFC GBC results across all pipelines

In a comparison of intra-PFC GBC results from all three preprocessing strategies, a difference between the MDD and HC groups was observed only with GSR. Specifically, a main effect for group with a cluster-defining uncorrected threshold of p<0.01, FWE-corrected at p<0.05, was found in the right superior frontal cortex (F=10.4, p<0.01, FWE-corrected, Figure 3 and Supplementary Table S7) and in the left middle frontal cortex (F=10.5, p<0.01, FWE-corrected). Individuals with MDD had significantly reduced intra-PFC GBCr at the baseline scan compared to HCs in the right superior frontal cortex (Z=3.7, p<0.001, FWE-corrected, *d*=1.6) and in the right middle frontal cortex (z=3.6, p<0.001, FWE-corrected, *d*=1.47; see Figure 3A and 3C). No significant main effects were observed for scan or for the interaction between group and scan (all p>0.05, FWE-corrected). As in previous studies [11,12], Fz-scores calculated with whole brain GBCr were extracted from the PFC GM mask and the values were compared between groups. In MDD participants, a small effect was seen for reduction in total PFC Fz-scores (mean F_Z_=0.0387, SD=0.0387 compared to HCs (mean: F_Z_=0.0405, SD=0.009, t=7, p<0.001, effect size for the difference: *d*=0.16), see Figure 3B).

**Figure 3.**
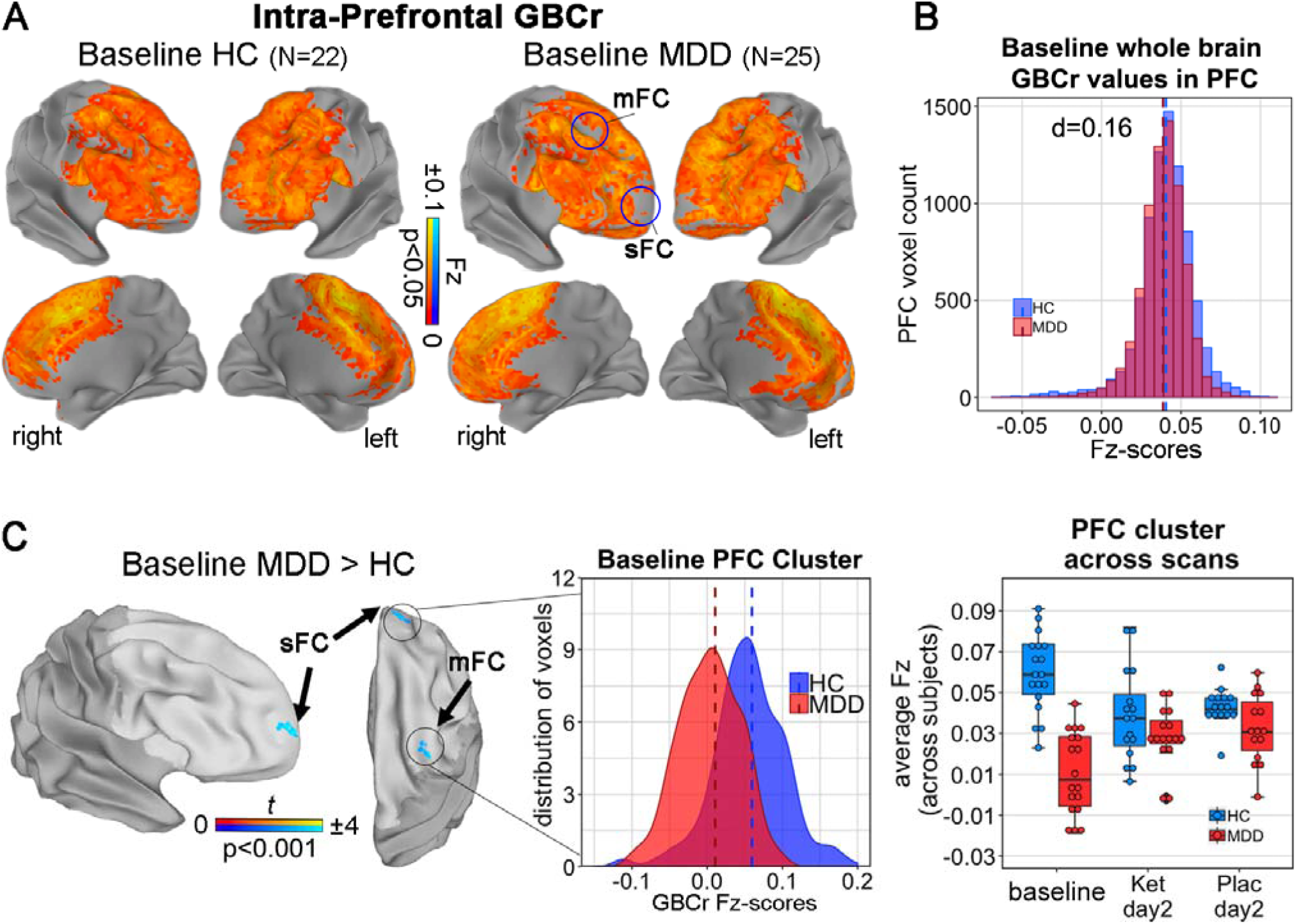
Reductions of intra-prefrontal cortex (PFC) global brain connectivity (GBC) with global signal regression (GSR; GBCr) in individuals with MDD. **A**, Average baseline GBCr in the PFC in both groups. Blue circles correspond to (**C**) and indicate reduced baseline GBCr in individuals with MDD. **B**, Baseline whole brain GBCr Fisher transformed Z-scores (Fz-scores) in the total PFC mask plotted for both groups indicating no difference, in contrast to previous studies (see Figure 3B in the study by Abdallah and colleagues, who reported a strong reduction with *d*=0.95 [12]). **C**, Reduced GBCr as indicated in (**A**) (p<0.05, FWE-corrected, initial p=0.001). As with total gray matter (GM) GBCr, no significant effect was seen versus baseline for ketamine, placebo, or treatment (all p>0.05, FWE-corrected). sFC, superior frontal cortex; mFC medial frontal cortex.

## Discussion

The principal objective of this study was to independently replicate previous findings of altered functional connectivity in individuals with MDD by recreating the preprocessing procedures used in previous work as rigorously as possible. In line with earlier results, group differences in GBC were detected between individuals with MDD and HCs only after using GSR. Contrary to our original hypothesis, PHYSIO regression did not produce results similar to those seen using GSR.

By systematically pursuing alternative preprocessing strategies, these data support several previous studies that used the same methodological approach [10-12,15,29,30]. In particular, reduced baseline connectivity was noted in MDD participants compared to HCs in the MCC and ACC as well as in the PFC in similar—though not identical—clusters, consistent with prior work and analytic methods [12]. Nevertheless, the results observed here in the MCC occurred in the opposite direction. Such replication attempts are vital to developing rsfMRI markers for MDD. In addition, GBCr was investigated in data drawn from randomized, double-blind, placebo-controlled ketamine trials. In contrast to previous studies [11,12,29] that did not use placebo controls and obtained scans closer to the time of infusion, no treatment effects on GBCr were observed.

### Replicating GBC alterations in MDD

The goal of this study was to replicate previous findings of regionally reduced network connectivity in individuals with MDD compared to HCs in the lateral PFC and superior medial frontal cortex, as well as increased GBCr in the cerebellum, precuneus posterior cingulate cortex (PCC), and lingual cortex [12]. However, in the present study, effects were only found after GSR calculated within the total GM. While significantly altered baseline GBCr was observed, our MCC cluster was located anterior to the PCC cluster reported by Abdallah and colleagues [12], and ours exhibited hypoconnectivity instead of hyperconnectivity. Given our rigorous statistical thresholds, the finding of PFC hypoconnectivity was not replicated, although it should be noted that at p<0.01 we found baseline hypoconnectivity in MDD participants to a similar spatial extent as in the bilateral ACC and medial superior/medial frontal cortices. Lateral PFC hypoconnectivity was not observed. In addition, the finding of reduced total PFC Fz-scores in MDD participants at baseline was not replicated (see Figure 3B vs. Figure 3 in Abdallah and colleagues [12]). Rather, our use of intra-PFC GBCr—which was equivalent to the whole-brain GBCr used in earlier studies [16,29]—detected reduced baseline GBCr in the superior and middle frontal cortex that exhibited no visual overlap with results reported by Abdallah and colleagues [12].

It is worth noting that in the study by Abdallah and colleagues [12], frontal GBCr Fz-scores increased significantly post-ketamine compared to baseline (lateral PFC pre-ketamine: -0.04 vs post-ketamine: 0.39), while cerebellar Fz-scores remained unchanged. In contrast to several previous studies [11,12,16], the present study did not replicate any previously observed treatment effects of ketamine versus either baseline or placebo on GBCr in either MDD participants or in HCs. This discrepancy could be due to the longer scan interval post-ketamine (48-72 hours in the present study versus 0 or 24 hours in the study by Abdallah and colleagues [12]), but little is known about the temporal reactivity of GBCr in pharmacological fMRI. If GBCr changes following clinical response lasted from hours to at least three to seven days post-ketamine, such changes in GBCr would have been detected.

Basic neuroscientific studies have consistently identified synaptic loss, reduced dendritic complexity, and reduced neuron and glial cell density in the PFC of individuals with MDD [31,32]. Meta-analytic structural MRI and fMRI studies have corroborated findings of reduced volumes and dysconnectivity in the PFC of MDD participants [5,7]. In this context, it is most likely that study-related or subject-related variables could account for the differential findings in whole brain GBCr observed here. First, a possible explanation is that sequence parameters were different in our study compared to previous GBC studies, which could have impaired comparability, although rsfMRI has been demonstrated to be reliable between scanner types and coil channel numbers [33]. Second, and in contrast to previous work [12], our participants had their eyes closed, which significantly affects network connectivity [34]. This could explain the divergent findings and, potentially, the opposite directionality [34]. Third, FreeSurfer was used to segment structural MRIs, and AFNI’s afni_procy.py script was used to preprocess rsfMRI data; in contrast, previous studies used FreeSurfer and Matlab.

Recent studies have noted considerable differences between imaging software packages for task-based fMRI [35], and this could have impaired the comparability of the studies. Finally, sample-related variables may have accounted for the observed differences. While variables such as baseline MADRS score and number of previous antidepressant trials did not differ between the present study and that of Abdallah and colleagues [12], MDD participants in the latter study were on average 9.4 years older, had 20% fewer females, and had been sick for six fewer years. Although controlling for age, sex, and BMI did not substantially alter our results, the differences could nevertheless have impaired external validity. Future studies of clinical imaging markers should consider ways to standardize clinically relevant variables as well as sequence differences.

### Changes in GBC were found only after GSR

A novel aspect of this study was the systematic investigation of three different preprocessing strategies. When averaged over total GM voxels, PHYSIO regression explained less variance and was topologically dissimilar to GSR, even though cardiac and respiratory signals share variance with the GS [36]. GBCr Fz-scores centered around zero, resulting in GBCr maps with reduced Fz-scores, including previously reported negative correlations [17]. Thus, GSR seemed to affect voxels in a non-linear pattern—that is, a voxel with high Fz-scores before GSR did not necessarily exhibit high Fz-scores after it. Consistent with previous work [37], this study demonstrated that the R^2^ GS also overlapped with the venous system, a pattern not observed for either BASE or PHYSIO regression (see Supplementary Figure S5). The GS has many non-neuronal components, such as vascular reactivity, motion, or CO_2_ concentration that warrant removal [2], but it also contains neuronal components related to slow fluctuations in gamma-range local field potential power [38] and neuropsychological variables such as vigilance or arousal, which would be undesirable to remove from fMRI signals [39]. Notably, topological GS alterations have been observed in individuals with schizophrenia [40], suggesting disease-inherent components of the GS.

Correspondingly, we demonstrated regionally reduced variance in the lingual gyrus of individuals with MDD. As regards the neurocognitive group-specific fMRI analyses, rsfMRI data preprocessed with GSR exhibited increased R^2^ in neurocognitive tests [41]. Given that significant group differences in the present study were only observed after removal of the GS, further investigations into components of the GS that characterize individuals with MDD are necessary. Such a component could improve noise removal procedures in future imaging marker studies.

In addition, regionally-altered differences in R^2^ were observed across scans, both for the GS as well as for the BASE regression, with more absolute R^2^ at baseline scans. These findings may be due to neuronal as well as non-neuronal components of the GS. Non-neuronal noise may also account for WM/CSF, since these signals contain similar spurious signals from vasculature, respiration, and motion- or scanner-related artifacts [17]. Nevertheless, the significantly different R^2^ across scans, as observed for GS and BASE, raises key questions regarding pharmacological connectivity in individuals with MDD. How stable is GSR across scans in individuals with MDD? Could habituation effects in post-treatment scans be mediated by GS components such as vigilance and arousal?

## Study strengths and limitations

The placebo control, crossover design, and inclusion of HCs are all significant strengths of the present analysis, as is the systematic investigation of three different preprocessing strategies.

A limiting factor associated with this study is that data were drawn from an existing dataset. Thus, parameters such as scan interval, echo planar image (EPI) sequence, and whether eyes were open versus closed could not be changed to exactly match the settings of comparator studies. Furthermore, the impact of scanner, sequence-related variables, test-retest reliability, and population-based variables on GBCr could not be assessed. The relatively small sample size is another limitation.

## Conclusions

The current study investigated the impact of preprocessing strategies on previously published GBC results and found that they depend on the use of GSR in preprocessing. At baseline, the present study detected altered connectivity in the cingulate and frontal cortices of individuals with severe MDD. However, in contrast to previous studies [11,12], hypoconnectivity instead of hyperconnectivity was observed in the MCC and a different neuroanatomical pattern within the PFC. Furthermore, previously observed increases in connectivity after antidepressant treatment with ketamine were not replicated. Longer post-treatment scan interval, scanner, or subject-related variables may account for these discrepancies. The findings suggest that additional studies exploring preprocessing levels are warranted before costly trials with experimental drugs are begun. Although rsfMRI analyses such as GBCr are being developed as candidate markers, the present study suggests that additional work is needed before they can be used in participants with mood disorders. In addition, future studies may wish to investigate which components of the GS may be specific to individuals with MDD.

## Funding and Disclosures

Funding for this work was supported by the Intramural Research Program at the National Institute of Mental Health, National Institutes of Health (IRP-NIMH-NIH; ZIAMH002857; NCT00088699), by a NARSAD Independent Investigator Award to Dr. Zarate, and by a Brain and Behavior Mood Disorders Research Award to Dr. Zarate.

Dr. Zarate is listed as a co-inventor on a patent for the use of ketamine in major depression and suicidal ideation; as a co-inventor on a patent for the use of (2*R*,6*R*)- hydroxynorketamine, (*S*)-dehydronorketamine, and other stereoisomeric dehydro and hydroxylated metabolites of (*R,S*)-ketamine metabolites in the treatment of depression and neuropathic pain; and as a co-inventor on a patent application for the use of (2*R*,6*R*)-hydroxynorketamine and (2*S*,6*S*)-hydroxynorketamine in the treatment of depression, anxiety, anhedonia, suicidal ideation, and post-traumatic stress disorders. He has assigned his patent rights to the U.S. government but will share a percentage of any royalties that may be received by the government. Dr. Kraus received travel support from Roche and AOP Orphan. All other authors have no conflict of interest to disclose, financial or otherwise.

## Data Availability

All data and code are available from Dr. Carlos Zarate upon request.

## Acknowledgements

The authors thank the 7SE research unit and staff for their support. Ioline Henter (NIMH) provided invaluable editorial assistance. Preprocessing of all rsfMRI data was conducted on linux CentOs at the NIH’s High-Performance Computing Biowulf cluster (http://hpc.nih.gov).

## Notes

### Clinical Trial

NCT00088699

